# Functional and Metabolic Adaptations of Blood Flow Restriction Exercise in Rats and Post-surgical Patients

**DOI:** 10.64898/2026.05.26.26353908

**Authors:** Andin Fosam, Susana Castelo Branco Ramos Nakandakari, Matthew Dworkowitz, Zongyu Li, Christopher Petrosino, Christopher Taber, Christina R. Allen, Rachel J. Perry

## Abstract

Blood flow restriction exercise (BFR-E) has gained popularity as a therapy used to improve muscle mass and strength in various clinical populations. However, the systemic and intramuscular responses to BFR-E are widely unknown. Here, we describe the functional and metabolic responses to BFR-E in our novel *in vivo* method of BFR-E in rats. Implementation of the model revealed increase in muscle mass and maximal strength in rats exposed to chronic BFR-E. Systemic metabolites related to glycolysis and redox metabolism were altered following acute BFR-E and metabolites related to amino acids and the TCA cycle were altered following chronic BFR-E. Moreover, transcriptomic analysis revealed a muscle-specific metabolic response to chronic BFR-E that coincided with morphological adaptations. Given the broad application of BFR-E in the rehabilitative setting, we examined the acute, systemic response to BFR-E in post-surgical human subjects. Semi-targeted metabolomic analysis revealed no significant alterations in circulating metabolites following acute BFR-E in humans, mirroring findings in acutely exercised rats. Together, these results suggest that the benefits of BFR-E are not mediated by acute systemic metabolic perturbations but instead arise from tissue-specific adaptations that develop with repeated exposure, establishing a conserved, translational framework for mechanistic investigation.

## Introduction

BFR-E is an increasingly utilized therapy to improve muscle mass and strength in recreational and rehabilitative settings for a range of individuals (1–3). Over the past two decades, several studies have demonstrated the efficacy of blood flow restriction exercise (BFR-E) to enhance muscle function, as it relates to sports performance and recovery from musculoskeletal injury (1,4–8). Seemingly paradoxically, BFR-E induces positive functional outcomes despite utilizing loads significantly below the recommended stimulus to induce muscular adaptation during resistance exercise (10-20 percent of an individual’s one repetition maximum versus 60-80%) (9–11). Robust discussion exists surrounding the underlying physiologic drivers of this phenotype (12–15). However, this discussion primarily relies on theoretical speculation, as a comprehensive physiological characterization of the systemic and intramuscular response to BFR-E is not well developed. Here, we address this gap by integrating functional, metabolic, and structural responses to BFR-E in animal and human models.

Muscle hypertrophy and strengthening occur via external overload stimuli that induce mechanical tension, muscle damage, and metabolic stress, driving anabolic adaptation (16). Mechanical tension, via force generation and muscle body stretch, causes perturbations to normal skeletal muscle architecture, wherein the muscle must structurally adapt to a large functional demand (17). This adaptation leads to an increase in muscle fiber size and number of contractile units within the fiber, providing additional room and machinery for greater force output(18). Studies also suggest an anabolic role of metabolic stress in the setting of an accumulation of metabolites, hormones, myokines, and exerkines following resistance exercise (19–21). These factors act upstream of protein synthesis regulators like mammalian target of rapamycin (mTOR) and contribute to subsequent increases in muscle protein synthesis (22). How BFR-E relates to these frameworks of muscle hypertrophy and strengthening is not completely understood.

Existing theories postulate that BFR-E may accelerate muscle fatigue, mechanical tension and metabolic stress, potentially forcing anabolic adaptation at significantly lower loads (23). There is some evidence that localized intramuscular hypoxia increases reactive oxygen species (ROS) activity and lactate production (4,24,25), but data are limited. Structural changes are also proposed to contribute to muscle growth following BFR-E. As type 2 (fast) muscle fibers are highly recruited during resistance exercise, some studies suggest that BFR-E may induce type 1 fibers to transition to a more type 2-like phenotype (12). This may increase the capacity of oxygen-restricted muscles to more quickly meet energy demands. Robust physiologic data are crucial for contextualizing these theories and ultimately for constructing a mechanistic framework of BFR-E.

Rodent models of BFR-E have recently been utilized to investigate underlying BFR-E physiology (25–30). Initial findings demonstrate that BFR-E may enhance the expression of key regulators of glucose transport and mitochondrial biogenesis in the tibialis anterior muscles of rats (30) and upregulate downstream targets of mechanistic target of rapamycin (mTOR) (28). However, animals in these studies were subjected to BFR with electro-stimulated muscle contraction under anesthesia, which may not reflect the physiologic responses to BFR-E in awake, exercising animals. High-throughput profiling of BFR-E is limited, but transcriptomic analysis has identified upregulated protein turnover as an important regulatory factor of the muscle hypertrophy observed in the soleus of rats undergoing BFR alone (31). While these data contribute to our understanding of BFR-E physiology, there is still much to be uncovered.

Recognizing this gap, we recently developed a novel *in vivo* rodent model of BFR-E (32). Herein, application of this model, as described here, explores the functional, metabolic, and structural responses to self-motivated BFR-E in rats. We also examine the systemic responses to rehabilitative BFR-E in the post-surgical setting in humans. The findings detailed here offer a perspective of BFR-E physiology that lays a nuanced foundation for studies of BFR-E mechanisms.

## Methods

The authors recognize the importance of sex as a biological variable and have accounted for this by including male and female participants (all genders) in the human study of BFR-E. While the use of male and female rats is crucial to fully understand the physiologic and metabolic responses to BFR-E, animal experiments presented here were limited to male rats. First, male rats are generally larger, which facilitated experimental procedures like blood collection, BFR-cuff placement, and resistance exercise. Second, literature suggests that male rats elicit a larger muscle hypertrophy response following muscle loading exposure compared to female rats. The authors posited the use of male rats would allow better delineation of the systemic and intramuscular responses to BFR-E. Lastly, studies were restricted to male rats to limit hormonal fluctuations in female rats due to the estrous cycle. Conducting similar experiments in female rats is crucial to generating a robust framework of BFR-E that includes sex-specific differences.

### Animal Experiments

All animal experiments were performed in accordance with the protocols approved and renewed by the Yale University Animal Use and Care Committee (Protocols #2022- and 2025-20290, respectively). 200-250g male Sprague-Dawley rats were obtained from Charles River Laboratories and housed in a 12 hr light/dark cycle with *ad lib* access to food and water in our animal facility. Terminal studies were performed 6 and 10 weeks after arrival in our facility for acute and chronic protocols, respectively.

#### Resistance Exercise Training Protocol

Rats underwent hindlimb-isolated knee extension resistance either with or without blood flow restriction as previously described (32). In the acute condition, 8-12-week-old (∼300g) male rats were assigned to one of two experimental groups: low-weighted resistance exercise without BFR (Ex Ctrl) or low-weighted resistance exercise with BFR (BFR-E). Animals underwent a 7 to 12-day acclimation period, after which they underwent internal jugular vein catheterization and a subsequent recovery period, as previously described (32). Rats were re-acclimated for 3-4 days to avoid de-training effects, after which the animals performed a single session of knee extension exercise loaded to 30 percent of their body weight. Exercise sessions lasted 10 minutes. Animals achieved a single repetition by extending from the starting “Rest” position (full knee flexion) to the top position (full knee extension) and returning to the starting “Rest” position. Cardboard tubes and plastic tube racks were utilized as tools to motivate repetitions, while taking care to avoid compensatory work done by front limbs. Rats were then immediately euthanized and whole quadriceps muscles were isolated. The isolated tissue was immediately flash frozen and stored at -80°C for future analysis.

In the chronic condition, rats were assigned to either the Ex Ctrl, BFR-E, or non-exercised sedentary control (Sed Ctrl) group. Rats were exercised for six weeks (5 consecutive days per week) under loads equal to 30 percent of their body weight. Prior to the start of the training period, rats were acclimated to the exercise protocol for 7-12 days. For each training session, the total number of repetitions performed in 10 minutes was recorded. Non-exercised sedentary controls did not engage in any resistance exercise over the six training weeks following the acclimation period.

#### Functional Testing

Maximal squat load was used to evaluate muscle strength following chronic resistance exercise with or without BFR. Maximal squat testing was conducted following acclimation in all experimental groups, but prior to the start of the exercise training protocol. Post-training testing was performed 72 hours following the final training session in all experimental groups. To appropriately evaluate maximal strength, rats performed hindlimb knee extension exercise against progressively increasing load until volitional failure. The load progression occurred as follows: 180g, 200g, 220g, 240g, 260g, 280g, 300g, 320g, 330g, 340g, 350g, 360g, 370g, 380g, 390g, and 400g. Progression in loaded weight was dependent on the successful completion of one repetition of the previous load. Volitional failure was determined after three failed attempts to complete a full repetition.

#### Body Composition Testing

Body composition was measured prior to the start of exercise training (baseline) and at the end of each training week. Lean mass, fat mass, and total body water were collected using the EchoMRI Body Composition Analyzer (EchoMRI, Houston, Texas). Body weight was recorded using a precise digital scale.

#### Terminal Studies

One day following functional testing, rats were implanted with internal jugular vein catheters for stable venous blood collection. Rats recovered for four days following catheterization, after which they performed two consecutive days of resistance exercise sessions (10 minutes per session, loaded to 30% of body weight), to offset the effects of de-training following the catheterization procedure and subsequent recovery. The following morning, blood venous blood samples were collected, and rats were euthanized via pentobarbital injection. Vastus intermedius (VI) and rectus femoris (RF) muscles of the quadriceps were isolated for differential analysis of predominantly glycolytic and oxidative muscles following chronic BFR-E, respectively. These tissues were flash frozen and stored at -80°C for future analysis.

### Clinical Study

All clinical procedures were performed in accordance with the protocols approved by the Yale University Institutional Review Board (NCT05012982). All participants completed informed consent protocols.

#### Study Visits

Healthy adults aged 18 to 65 years who had undergone anterior cruciate ligament reconstruction (ACL-R) surgery were recruited to participate in two study visits that included weighted single-leg knee extension exercise with or without BFR (Supplemental Fig 1a). Participants were eligible if ACL-R surgery occurred at most 6-8 weeks prior to the first study visit. Exclusion criteria included a positive medical history of heart disease, lung disease, rheumatoid arthritis, diabetes mellitus, a history of one or more tears of the same ACL, sickle cell trait or disease, past or present smoking or tobacco use, recent infection, pregnancy, history of deep vein thrombosis, or medications that increase clotting risk. Healthy control participants were recruited within the same inclusion and exclusion criteria, except that healthy participants had no history of ACL-R or other musculoskeletal procedures or pathologies. Study visits were conducted either at the Yale New Haven Health Orthopaedic and Rehabilitation Clinic or the Gaylord Health Physical Therapy Clinic as approved by the Yale IRB.

#### Exercise Protocol

Study visits included single-leg knee extension exercise performed on a Total Gym Apparatus (Total Gym GTS Classic Gravity System, Total Gym, San Diego, USA) set to an incline that allowed an effort of approximatebhly equal to 30 percent of the participant’s body weight. Participants performed a total of 90 repetitions on the surgical limb, as set by the following protocol: one set of 30 repetitions followed by four sets of 15 repetitions with 30 seconds of rest between sets. The exercising leg was chosen at random for healthy control participants. The maximal time allotted to complete the exercise protocol was seven minutes, after which the occlusion cuff was deflated and removed.

#### Blood Flow Restriction Parameters

The Delfi Cuff (Owens Recovery Science) was used to achieve and maintain blood flow restriction during the exercise protocol. Total limb occlusion pressure was determined for each participant immediately prior to the start of the exercise session for each study visit. The occlusion cuff was placed on the proximal thigh, as close to the hip joint as feasible. To achieve therapeutic BFR, cuff pressure was set to 70% of the individual’s total limb occlusion pressure. During the non-BFR control visit, cuff pressure was set to a sub-therapeutic 40% occlusion to mimic experimental conditions without inducing physiologic blood flow restriction.

#### Blood Collection

A 22G intravenous catheter was placed in the antecubital vein prior to the start of the exercise protocol. Whole blood was collected into EDTA-coated tubes (BD Vacutainer k2 EDTA, BD, Franklin Lakes, NJ) immediately prior to the exercise session, 30 minutes, and 60 minutes post-exercise. A small aliquot (100uL) of whole blood from each collection was mixed with 10uL of 0.5M EDTA in Eppendorf tubes pre-coated with 2M EDTA for complete blood count analysis. Blood samples were stored on ice during the study for further processing.

Within one hour of the final blood draw, blood samples were centrifuged, and plasma was collected into standard 1.5mL Eppendorf tubes. Plasma was stored at -20°C for future analysis.

#### Qualitative Surveys

Pre-exercise soreness surveys were administered to participants to establish a baseline level of perceived pain prior to each study visit. Following the exercise session, participants were asked to complete demographic and activity surveys, which collected information regarding habitual exercise behaviors. Soreness and tolerability surveys were also conducted on a 1-10 scale to evaluate post-exercise perceived pain and exercise difficulty (Supplemental Fig 1b,c).

### Biochemical Analyses

#### Immunological and Inflammatory Characterization- Rat and Humans

Prepared blood samples were stored at room temperature and measured within five hours of collection. Automated blood counts were acquired using a Hemavet 950FS Auto Blood Analyzer (Drew Scientific, Boston). Quantified blood markers included white blood cells, neutrophils, lymphocytes, basophils, eosinophils, monocytes, hemoglobin, and hematocrit. Inflammatory cytokine and chemokines were quantified using Discovery Assays available through Eve Technologies.

#### Quantitative Polymerase Chain Reaction Protocol- Rats

TRIzol was utilized to isolate total RNA from whole quadriceps (acute), rectus femoris and vastus intermedius (chronic) muscle, and an RNeasy kit (Qiagen) was used to further purify the RNA. Isolated RNA was converted to cDNA using the High-Capacity cDNA Mastermix (Applied Biosystems, Waltham, MA) and reverse transcribed using the T100 Thermal Cycler (BioRad, Hercules, CA). Gene expression was quantified using an Agilent Stratagene Mx3005P QPCR System (Agilent, Santa Clara, CA) and detected via the iTAq Universal SYBR Green Supermix (BioRad, Hercules, CA). A 1:10 dilution of each cDNA sample was prepared. Primers pairs for each gene of interest were diluted to a final concentration of 100 nM, except for *Hif1a* and *Myh2*, which were diluted to 50nM. Primer sequences are detailed in Supplemental Table 1. Each reaction contained 5ul of the SYBR Green Supermix, 0.5ul of each 100 nMol primer within the primer pair, 1 ul cDNA (1:10 dilution), and 3ul of RNA-ase free water to achieve a final volume of 10ul per well. Amplification reactions were performed in qPCR 96-Well plates (Agilent, Santa Clara, CA) within the following thermal parameters: 1 cycle (15 minutes at 95° C); 40 cycles (10 minutes at 95° C → 30 seconds at 60° C); 1 cycle (1 minute at 95° C → 30 seconds at 60° C → 30 seconds at 95° C). Each sample ran in duplicate and replicate Ct values obtained for each sample were averaged.

Rat hypoxanthanine phosphoribosyltransferase 1 (*Hprt1*) served as a housekeeping gene to normalize cDNA content in each tested sample. Expression of each target gene was evaluated relative to a non-exercised sedentary control group.

#### Immunohistochemical Staining- Rats

At the time of sacrifice, the contralateral whole quadriceps was collected from the animals, fixed in 10% formalin overnight, and then transferred to 70% ethanol until processing for immunohistochemistry. Muscles embedded in paraffin blocks, sectioned (5 microns) perpendicular to muscle fiber direction. Muscle sections were incubated in antibody against Fast Myosin (1:800 dilution, MY-32, Sigma) and slow Myosin (1:200 dilution, AB234431, Abcam). Fast Myosin was then labeled with anti-rabbit HRP-conjugated secondary antibody and detected with diaminoebzidine, while slow myosin was labeled using anti-rabbit alkaline phosphate and detected with fast red. Whole tissue scans were acquired at 20x magnification using the Motic Easy Scan One (Motic, Emeryville, CA).

#### Muscle Fiber Count and Typing- Rats

Whole tissue scans were divided into eight regions from which mages were captured at 10x magnification. Muscle fiber number and cross-sectional area were determined using Fiji software. Fast and slow muscle fibers were identified by visual inspection of fibers that were stained brown (fast) or pink (slow) (Supplemental Fig 3). Approximately, 1100 muscle fibers were analyzed per sample.

#### Metabolomics Sample Preparation, Extraction, and Quantification- Rats and Humans

For metabolomics analysis, blood samples were collected from rats following terminal experiments in the acute and chronic conditions. Whole blood was centrifuged and plasma was extracted and stored at -80° C until analysis.

Metabolites were extracted by adding 30 μl of extraction solution A (50:50 acetonitrile/methanol) to 10 μl of plasma. Samples were vortexed for 15 minutes at 4° C and centrifuged at 21,000 × g for 15 minutes at 4 °C. The supernatant was then transferred to LC/MS vials. Metabolites were resolved on a Vanquish U-HPLC system coupled to a Q Exactive HF-X hybrid quadrupole-Orbitrap mass spectrometer (Thermo Fisher Scientific, Waltham, MA) equipped with a HESI source operating in negative ion mode. The analytes were separated on an iHILIC column (5 μm, 150 × 2.1 mm I.D., HILICON). The iHILIC column was equilibrated with the following buffers: A (water containing 20 mM ammonium carbonate and 0.1% ammonium hydroxide, pH 9.3) and B (acetonitrile). The Vanquish U-HPLC was operated at a flow rate of 0.150 ml/min with the following gradient: 0 - 23 min, a linear gradient from 95% B to 5% B; 23 - 25 min, held at 5% B; 25–25.5 min, returned to 95% B at 0.20 mL/min (to waste); 25.5–32.5 min, held at 95% B; and 32.5–33 min, 95% B at 0.15 mL/min. Ions were analyzed at a resolution of 60,000 at m/z 200 over a scan range of m/z 70 - 1000. The AGC target was set to 1e6, and the maximum injection time was 100 ms.

Targeted feature extraction and quantification of MS data were performed using Skyline software(33). Peak area integration and metabolite identification were accomplished based on accurate mass and retention time and were curated using an in-house library of standard compounds. Metabolite profiling and statistical analyses, including pairwise, were performed using Graphpad Prism software.

### Statistics

Graphpad Prism 10 was used for all statistical analyses. Data are presented as mean ± standard deviation unless otherwise specified. Differences between two groups were calculated by the 2-tailed unpaired Student’s *t*-test. Differences between two variables within the same animal were examined with a paired Student’s *t*-test. Differences between three groups were calculated using a one-way or two-way ANOVA with multiple comparisons. Bonferroni correctons were applied to t-test and ANOVA calculations. Statistical significance was set at p < 0.05. T statistics are reported as t (degree of freedom). To evaluate the practical significance of observed differences, both effect sizes and percentage changes (%Δ) were calculated. Cohen’s d were considered trivial, small, moderate, large, and very large when the values were 0-0.2, 0.2-0.6, 0.6-1.2, 1.2-2.0, and >2.0, respectively, based on the classification proposed by Hopkins (2002)(34).

## Results

### Rodent Study Results

Rats were acutely- and chronically-exercised as depicted in Fig 1a. There was no observed difference in body weight between groups (Fig 1c), but we found that acute BFR-E rats performed fewer repetitions in the allotted time compared to Ex Ctrl rats (Fig 1d).

**Figure 1.**
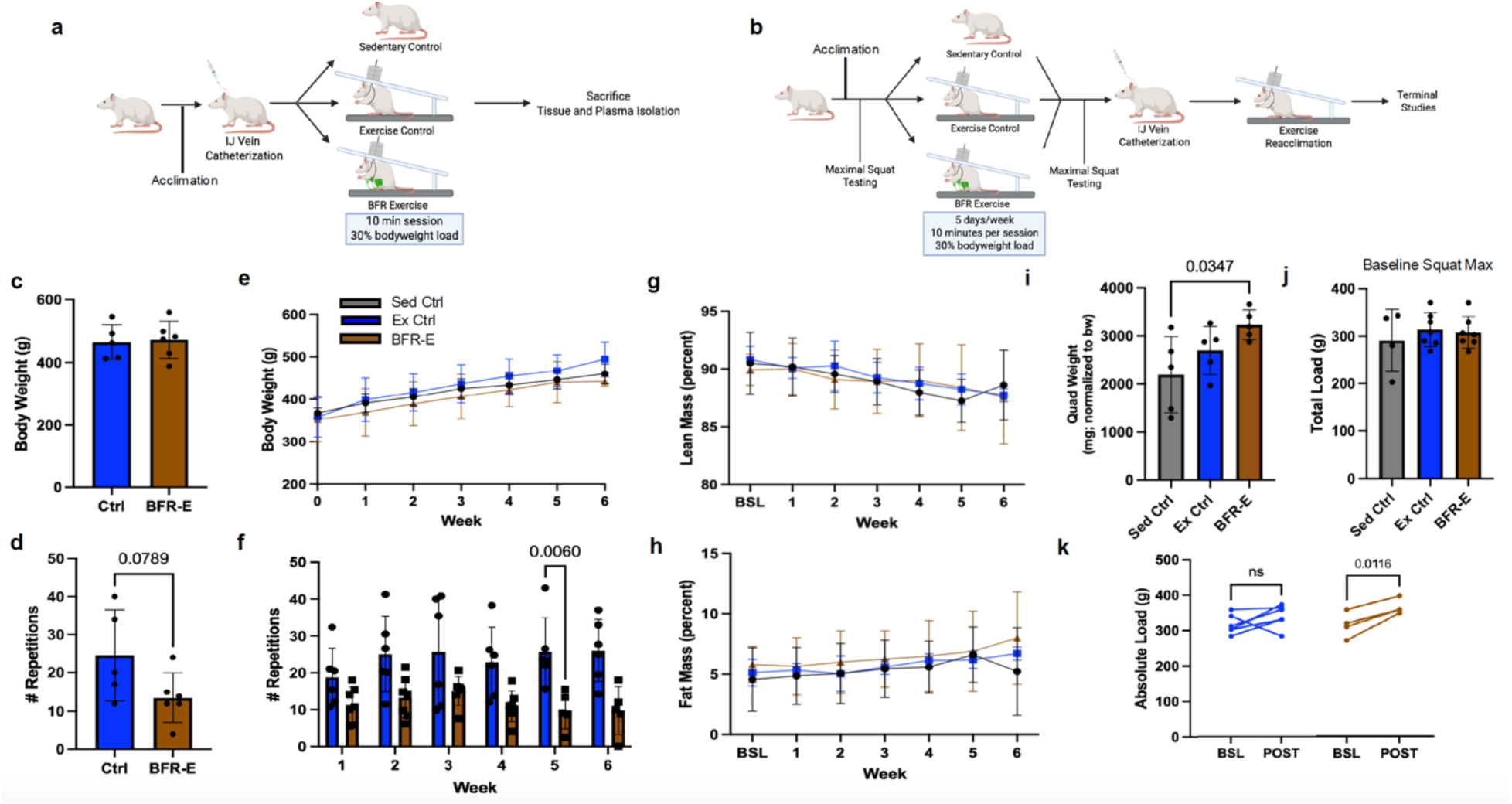
Effects of acute and chronic BFR-E on functional work, muscle hypertrophy, and muscle strengthening. Graphical schematic depicting the (a) acute and (b) chronic exercise protocols. (c) Body weight and (d) number of repetition differences between exercise control (blue) and BFR-E (brown) rats after a single session of resistance exercise. (e) Body weight and (b) number of repetition differences between sedentary (black), exercise control (blue) and BFR-E (brown) over six chronological weeks of resistance exercise training. Longitudinal comparison of (g) fat and (h) lean mass across six weeks of chronic resistance exercise. (i) Quadriceps weight comparison among experimental groups following chronic resistance exercise. (j) Baseline maximal squat strength prior to six weeks of resistance exercise. (k) Intra-animal comparison pre and post chronic resistance exercise maximal strength. *p<0.05 IJ = internal jugular; BFR-E= Blood flow restriction exercise; BSL = Baseline, Quad = Quadriceps; Sed Ctrl = Sedentary Control; Ex Ctrl = Exercise Control

Following six weeks of chronic exercise, BFR-E rats maintain body weight comparable to that of the sedentary and exercise control groups (Fig 1e). Compared to the exercise control, BFR-E rats performed fewer repetitions each week, despite statistical insignificance (Fig 2f).

**Figure 2.**
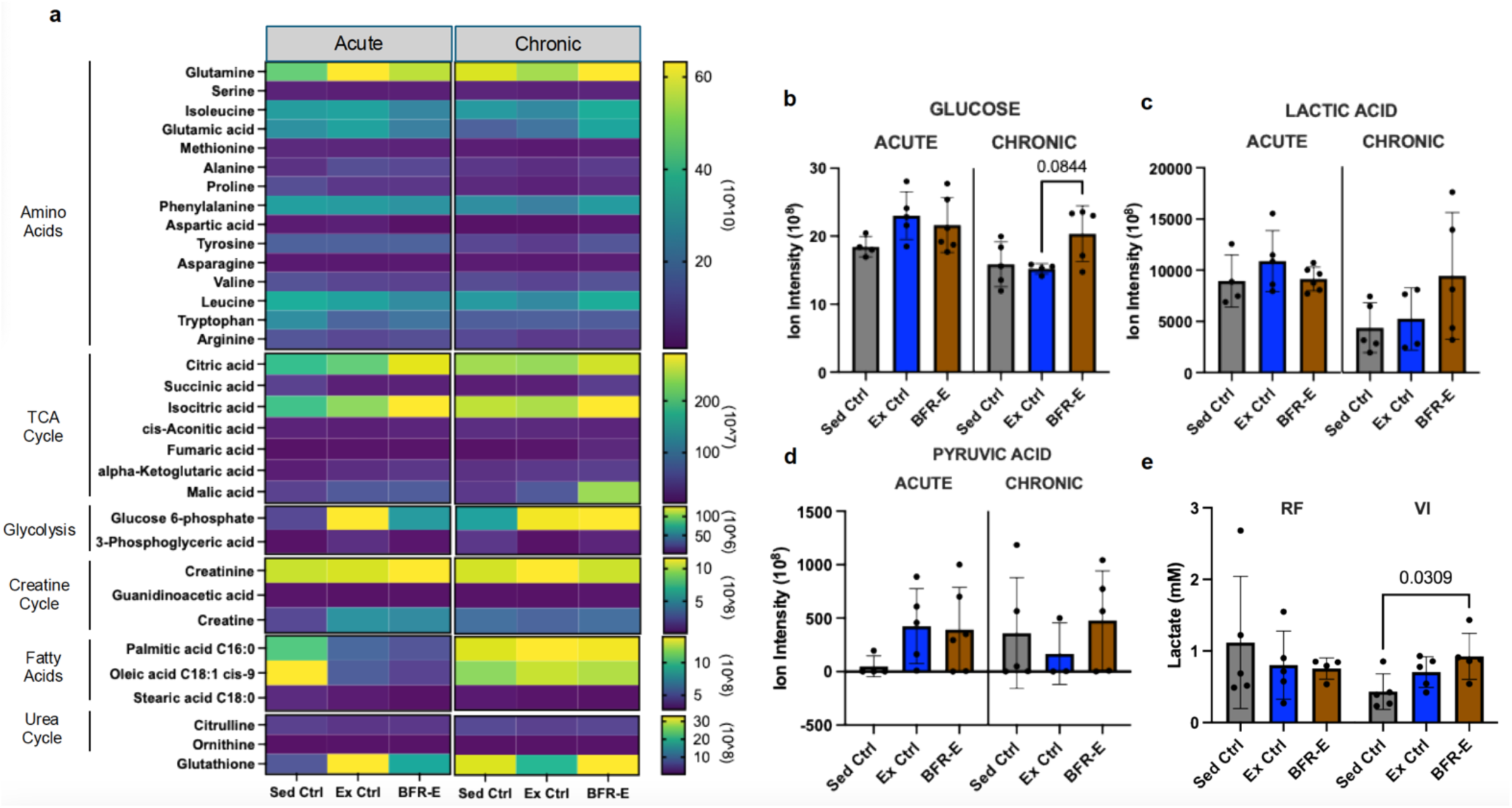
Metabolomic response to acute and chronic BFR-E in rat plasma. (a) Heatmap demonstrating intergroup comparisons of differential metabolites identified by semi-targeted metabolomic probe using liquid chromatography mass spectrometry analysis from isolated plasma in sedentary, exercise control, and BFR-E rats that underwent acute or chronic exercise. Data are presented as ion intensity. Yellow color indicates greater signal intensity, whereas purple color indicates lower signal intensity. (b) Bar graphs comparing ion intensities of glucose, (c) lactic acid, and (d) pyruvic acid from plasma of sedentary control, exercise control, and BR-E rats after acute or chronic exercise exposure. (e) Intramuscular lactic acid measured from the rectus femoris (RF) or vastus intermedius (VI) of rats that underwent chronic BFR-E. Data are presented as mean ± SD.

Body composition analysis revealed no difference in fat and lean mass (Fig 1g,h), however, BFR-E rats demonstrated a significant increase in quadriceps mass compared to the sedentary control group after chronic exercise training (Fig 1i). To evaluate the muscle-strengthening effects of BFR-E, maximal hindlimb strength was evaluated via single-repetition maximal squat testing. Prior to exercise training, maximal squat load was similar among all groups, indicating an equal strength level (Fig 1j). Following chronic exercise, the BFR-E group demonstrated a significant increase in maximal squat load, whereas sedentary and exercise control groups saw no significant change in squat load (Fig 1k).

These functional data suggest BFR-E to be a distinct stimulus to low-loaded exercise, which led to exploration of the broad systemic responses to BFR-E as it relates to the circulating metabolome. Thirty-three metabolites related to amino acids, citric acid cycle, glycolysis, creatine cycle, fatty acids, urea cycle, and redox reactions were probed from the plasma of acutely and chronically-exercised rats in each experimental group (Fig 2a). Several inter-group differences were observed in the acute and chronic conditions. Of note, acute BFR-E rats demonstrated significant decreases in signal intensity of glucose-6-phosphate, palmitic acid, oleic acid, and glutathione (Supplemental Table 2). Chronic BFR-E rats demonstrated significant increases in signal intensity of glutamic and malic acid. Further, there was an observed trending increase in plasma glucose following chronic BFR-E (Fig 2b), with no significant change in plasma lactic or pyruvic acid (Fig 2c, d).

We then investigated the intramuscular responses to BFR-E to explore the tissue-specific response of BFR-E to transient vascular restriction. Intramuscular lactate was measured in the Rectus Femoris (glycolytic) and Vastus Intermedius (oxidative) of chronically-exercised rats. Concentrations were similar across groups in the Rectus Femoris (RF), but significantly increased in the Vastus Intermedius (VI) of BFR-E animals when compared to sedentary control rats (Fig 2e). Hypoxia inducible factor 1a (*Hif1a)* gene expression was measured in whole quadriceps of acutely-exercised rats and in the RF and VI of chronically-exercised rats. Acute BFR-E does not alter quadriceps *Hif1a* gene expression compared to the sedentary and exercise control groups (Fig 3a). In the chronic condition, no difference in *Hif1a* gene expression among groups was observed in the RF (Fig 3h); however, *Hif1a* was significantly increased in the VI of BFR-E animals compared to the sedentary and exercise groups (Fig 3o).

**Figure 3.**
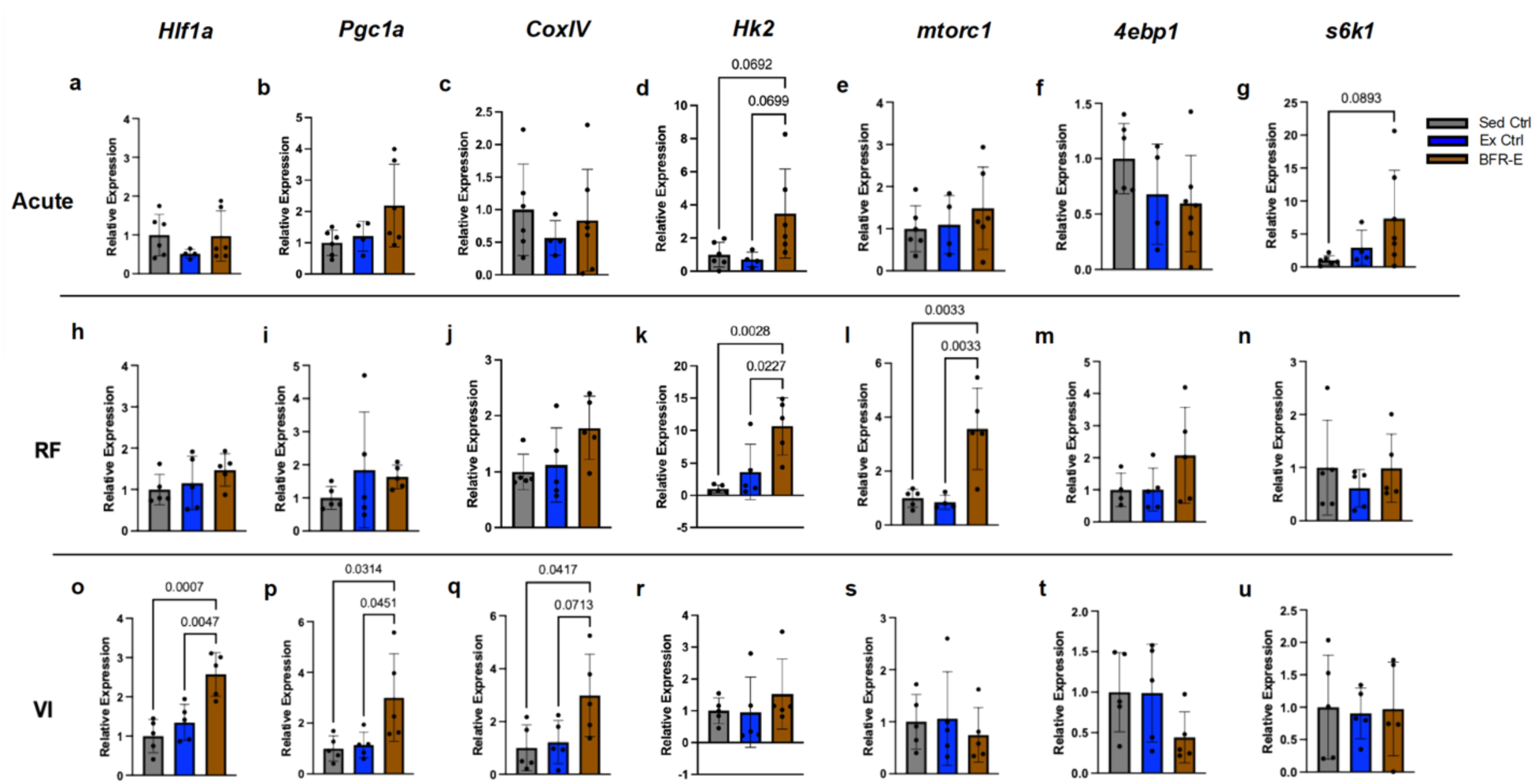
Acute and Chronic BFR-E increases intramuscular gene expression in muscle-specific fashion. Expression of genes related to hypoxia (*Hif1a)*, oxidative metabolism (*Pgc1a*, *CoxIV)*, glycolytic metabolism (*Hk2)*, and muscle protein synthesis (*Mtorc1*, *4ebp1*, *S6k1)* in (a-g) whole quadriceps of acutely exercised rats, (h-n) rectus femoris (RF) of chronically exercised rats, and (o-u) vastus intermedius (VI) of chronically exercised rats. *p<0.05

To investigate the effects of BFR-E on genes related to glycolytic and oxidative metabolism, we measured the gene expression of several key metabolites. We found no difference between groups in the intramuscular gene expression of peroxisome proliferator-activated receptor gamma co-activator 1-alpha (*Pgc1a*) or complex IV (*CoxIV*) in the acute condition (Fig 3b,c) or in the RF of chronic RE rats (Fig 3i,j). However, *Pgc1a* and *CoxIV* were significantly increased in the VI of chronic BFR-E animals (Fig 3p,q). Hexokinase-2 (*Hk2*) gene expression was measured to examine BFR-E effect on glycolytic metabolism in acute and chronic RE conditions. *Hk2* was similar between groups in the acute RE condition (Fig 3d), although expression in quadriceps of BFR-E rats was slightly increased compared to the control groups, with trending significance. *Hk2* gene expression in the RF was increased in the chronic BFR-E group (Fig 3k); however, this increase was not observed in the VI (Fig 3r).

Gene expression of mTOR and its downstream targets were quantified to determine the effects of BFR on protein signaling pathways. No intergroup differences were observed except for a trending increase in S6k1 gene expression in quadriceps of acute BFR-E rats (Fig 3e-g). mTOR gene expression was significantly increased in the RF of chronic BFR-E rats with no change in expression of downstream targets (Fig 3l-n). *mTOR*, *4ebp1*, and *S6k1* gene expression were comparable across groups in the VI of chronically-exercised rats (Fig 3s-u).

To assess structural changes to the quadriceps muscle following chronic BFR-E, muscle fibers were counted and characterized in each animal. We observed a pattern of decreasing muscle fiber number and increasing muscle fiber size in the BFR-E and exercise control groups compared to the sedentary control, however these observations were not statistically significant (Fig 4a-c). We observed a similar muscle fiber type distribution in the whole quadriceps of rats in each experimental group, with around 90% of fibers identified as fast (Fig 4d).

**Figure 4.**
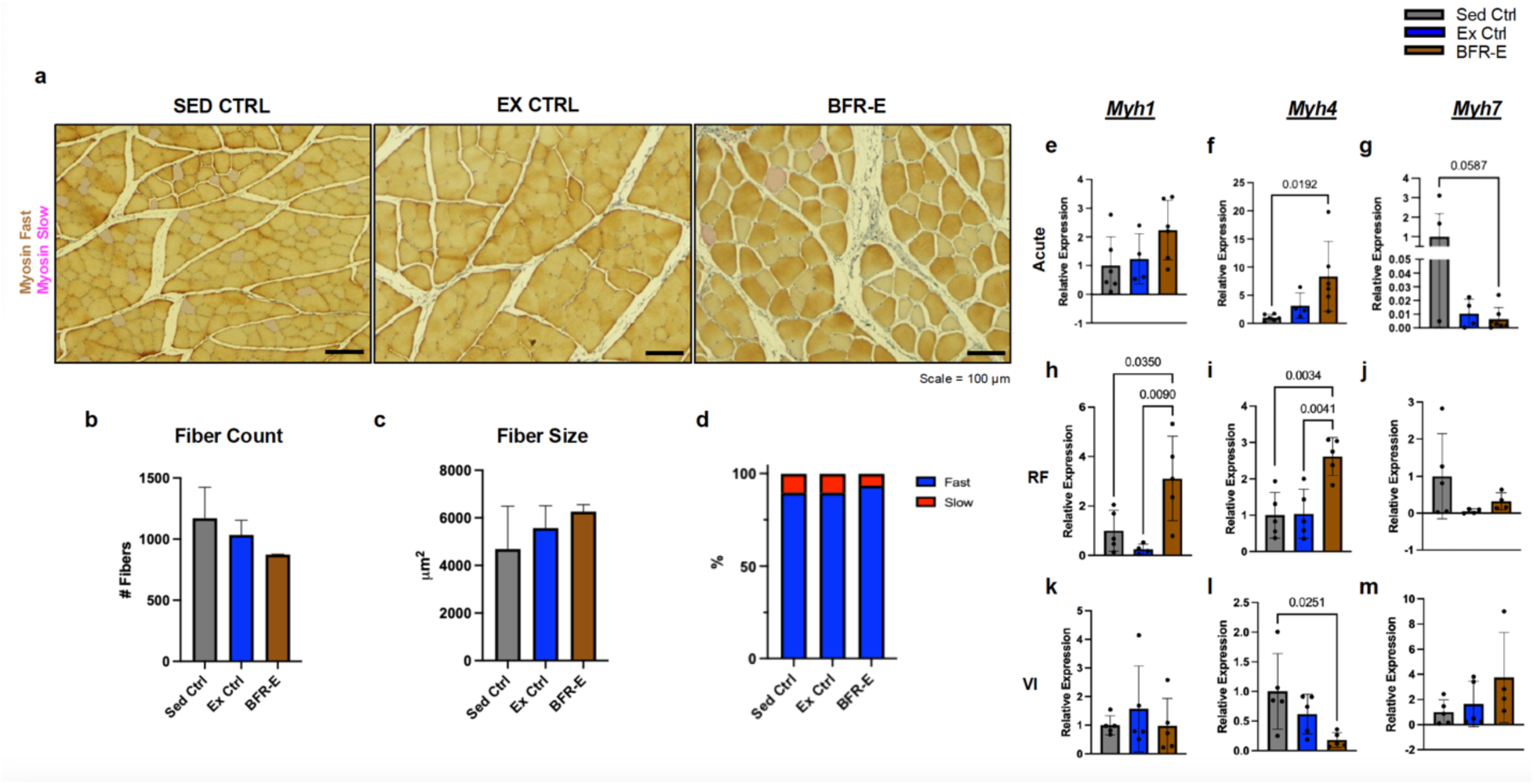
The effect of BFR-E on muscle morphology and fiber type recruitment in rats. (a) Representative cross-sectional images of the quadriceps muscle isolated from sedentary control, exercise control, and BFR exercise rats The staining method facilitated identification of fast (brown) and slow (pink) fibers. Images captured at 10x magnification. (b) Bar graphs demonstrating muscle fiber number and (c) muscle fiber size in sedentary (gray bar), exercise control (blue bar) and BFR-E (brown bar) rats. (d) Stack bar graph depicting the total proportion of fast and slow muscle fibers identified in whole quadriceps of rats in each experimental group. (e-g) Expression of genes encoding structural proteins for fast (*Myh1* and *Myh4*) and slow (*Myh7*) muscle fibers in whole quadriceps from acutely-exercised, (h-j) the rectus femoris, and (k-m) the vastus intermedius from chronically-exercised rats. Data are presented as mean± SD. Significance denotes as P<0.05.

To understand the molecular landscape underlying the potential structural changes observed following BFR-E, gene expression of myosin heavy chains encoding fast and slow structural proteins was quantified. Acute BFR-E led to an increase in fast type (*Myh4*) and a decrease in slow type (*Myh7*) fiber gene expression, with no significant change in Myh1 gene expression (Fig 4e-g). Chronic BFR-E rats demonstrated significantly higher expression of *Myh1* and *Myh4* in the RF compared to the sedentary control and exercise control groups (Fig 4h,i). No significant change in *Myh7* was observed in the RF, however there was a trending decrease in gene expression in the BFR-E and exercise control groups (Fig 4j). In the VI, *Myh4* was significantly decreased in the BFR-E group, with comparable expression levels of *Myh1* compared to the sedentary and exercise controls (Fig 4k,l). *Myh7* in the vastus intermedius was unchanged among groups, although a visual increase in expression was observed in the BFR-E group (Fig 4m).

### Human Study Results

Given the extensive evidence of the functional benefit of BFR-E in rehabilitative settings, we were eager to investigate the systemic responses to acute BFR-E in the post-surgical condition. Seven ACL-R participants and 11 healthy controls were recruited for two study visits that included a single session of single-leg knee extension exercise with or without BFR. Table 1 reports the subject characteristics. Age, sex, and the number of days between visits were similar between groups. Qualitative surveys were used to determine baseline activity levels. There was no difference between groups in the number of days per week or the number of hours per session spent engaging in strength or aerobic activity. Perceived pain was quantified using a soreness scale that ranged from 1 (no soreness) to 10 (severe soreness). We observed no significant difference in perceived pain between groups before and after the exercise session with BFR. However, ACL participants reported more soreness before the non-BFR exercise session. Exercise difficulty was determined using a rate of perceived exertion (RPE) scale to compare the tolerability of exercise with and without BFR. There was no difference in the tolerability of exercise with and without BFR between groups.

**Table 1.** Subject Characteristics Table. Demographic, activity, and exercise parameters in ACL and non-surgical control participants. Data are presented as mean ± SD.

|  | Control | ACL | p-value |
| --- | --- | --- | --- |
| N | 11 | 7 |  |
| Age (years) | 37.1 ± 10 | 39.5 ± 9.6 | 0.63 |
| Sex (% female) | 45.5 | 29 |  |
| Race/Ethnicity (NHW/HW/AA/A) | 5/2/2/2 | 3/2/2/0 |  |
| **Visit Parameters** |  |  |  |
| Time Between visits (days) | 16.8 ± 4.9 | 14.6 ± 3.0 | 0.3 |
| **Activity Parameters** |  |  |  |
| Strength Training (days) | 2.3 ± 2.2 | 2 ± 2 | 0.79 |
| Aerobic Training (days) | 4.2 ± 2.0 | 4.3 ± 1.4 | 0.91 |
| Strength Training (hours per session) | 1.4 ± 1.4 | 0.8 ± 0.5 | 0.4 |
| Aerobic Training (hours per session) | 0.78 ± 0.51 | 0.91 ± 0.2 | 0.54 |
| **BFR Parameters** |  |  |  |
| Limb Occlusion Pressure (mmHg) | 160 ± 17.1 | 168 ± 22.1 | 0.37 |
| Soreness Pre: non-BFR | 0.1 ± 0.3 | 1.00 ± 1.2 | **0.032** |
| Soreness Post: non-BFR | 0.8 ± 0.9 | 1.00 ± 0.8 | 0.65 |
| Soreness Pre: BFR | 0.72 ± 1.0 | 1.00 ± 0.6 | 0.53 |
| Soreness Post: BFR | 0.91 ± 1.4 | 1.7 ± 2.0 | 0.32 |
| Tolerability/RPE: Non-BFR | 1.9 ± 1.4 | 2.3 ± 1.6 | 0.86 |
| Tolerability/RPE: BFR | 3.09 ± 2.2 | 3.93 ± 1.9 | 0.42 |

Data presented as arithmetic mean ± SD; n= number of subjects. Abbreviations: ACL, anterior cruciate ligament; ; NHW, non-Hispanic white; HW, Hispanic white; AA, African American; A, Asian; BFR, blood flow restriction; RPE, rate of perceived exertion.

To better understand BFR-driven alterations to circulating metabolites related to amino acid, citric acid cycle, glycolysis, creatine metabolism, fatty acids, urea cycle, and redox metabolism, we performed a semi-targeted metabolomic probe of plasma acquired from ACL-R and control participants prior to, 30, and 60 minutes post the BFR-E study visit. We observed no time-dependent or group-dependent difference in circulating amino acids or metabolites related to TCA cycle intermediates, glycolysis, fatty acids, creatine cycle, urea cycle, or redox metabolism (Fig 5a). No inter-group or time-dependent differences were observed in the signal intensities of glucose, lactic acid, pyruvic acid, or creatine (Fig 5b-e). Collectively, these findings demonstrate that acute BFR-E does not elicit detectable systemic metabolic perturbations in either ACL-R participants or healthy controls. In the context of the well-established functional benefits of BFR-E, these data suggest that its efficacy is unlikely to be mediated by large, transient changes in circulating metabolites, but rather may arise from cumulative responses to local intramuscular adaptations to BFR-E.

**Figure 5.**
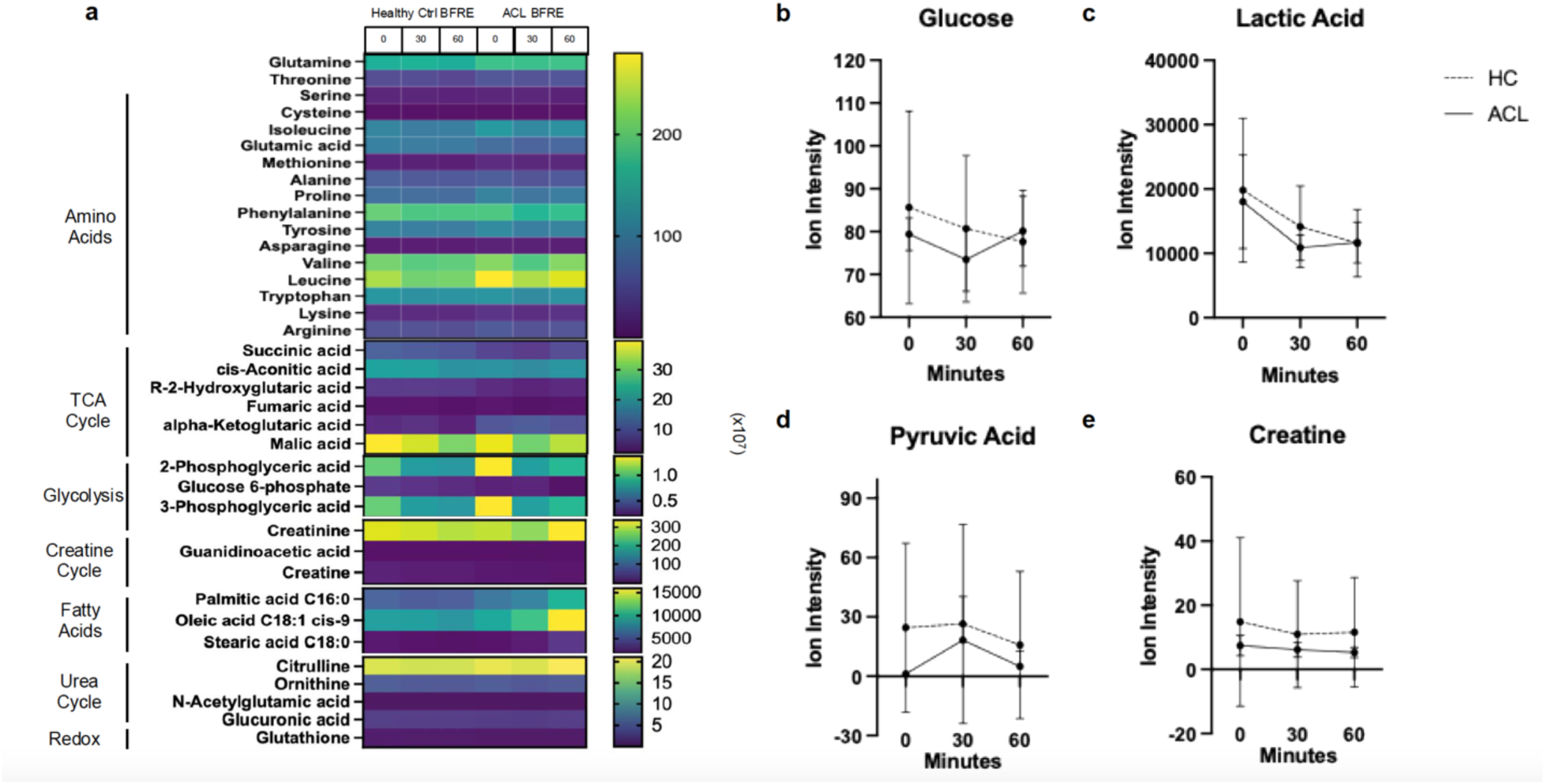
Metabolomic response to acute BFR-E in human plasma. (a) Heatmap demonstrating intergroup comparisons of differential metabolites identified by semi-targeted metabolomic probe using liquid chromatography mass spectrometry analysis in isolated plasma prior to exercise (0), 30 minutes (30) and 60 minutes (60) following a single session of BFR exercise from post-anterior cruciate ligament reconstruction and healthy non-surgical controls. Data are presented as ion intensity. Yellow color indicates greater signal intensity, whereas purple color indicates lower signal intensity. (b) Intergroup (ACL subjects are represented as dashed lines and healthy controls are represented as solid lines) and time-dependent comparisons of ion intensities of glucose, (c) lactic acid, (d) pyruvic acid, (e) and creatine quantified from plasma after acute BFR exercise exposure. Data are presented as mean ± SD.

## Discussion and Conclusions

Together, these data detail functional and molecular responses to acute and chronic BFR-E in rodents and systemic molecular responses to acute BFR-E in post-surgical patients. We previously described a novel *in vivo* model of BFR-E, which successfully induces and maintains BFR above the therapeutic threshold (>40% occlusion) and achieves weight-progressive knee extension exercise in rats (32). Upon implementation of this model, we show that BFR-E trained rats demonstrate increased muscle size and strength, despite performing less functional work. These data suggest that chronic BFR-E in rats induces muscle fiber-specific metabolic changes, which coincide with structural intra-muscular changes such as increased muscle size and muscle-specific enhancement of fiber type gene expression. We found minimal changes in circulating biomarkers and metabolites in post-surgical patients who underwent an acute session of BFR-E.

The positive functional change we observe in our rodent model of BFR-E (i.e. muscle hypertrophy and strengthening) provides a key basis for the physiologic and mechanistic understanding of BFR-E. These findings are especially noteworthy as BFR-E rats performed less functional work, despite observing significant and disparate muscle-specific responses. So, in our model, while BFR-E rats are exposed to roughly half of the external stimulus, they can achieve similar if not greater functional improvements, which our data suggest is physiologically distinct from non-BFR low-loaded resistance exercise. The nature of this reduced work and its relation to physiologic responses warrant further investigation. BFR-E rats performing fewer repetitions may be due to increased discomfort caused by the inflated cuff. However, our implementation of this novel model supports the evidence that BFR-E is tolerable across model systems. In our human study, we evaluated the effect of BFR-E on perceived pain. In our participants, pain scores were comparable between groups. While it was not feasible to directly examine BFR-induced discomfort in animals, BFR-E rats completed six weeks of resistance exercise with BFR and could tolerate the full duration of each session. Further, there were no observed functional deficits or signs of discomfort, including reduced activity, social isolation, or limping.

Other models of maximal strength testing in rats generally involve force-sensing grip strength meters of progressively-weighted ladder climbing(35–38). While these models test maximal output, they either are not standardized and/or cannot be applied to the evaluation of BFR functional outcomes. For instance, grip meters specifically test forelimb strength. BFR has not yet been applied to rodent forearms. Weighted ladder climbing can be progressively loaded, but involves the contribution of un-occluded limbs, introducing potential compensation. Here, we adapted use of the squat apparatus described previously to test maximal strength (32). Briefly, this apparatus uses a long lever arm with a flexible harness attached on one end for placement of the animal. The arm can be stacked with weights as desired. To test maximal strength as it relates to BFR-E, we designed and employed a weight-progressive single repetition schema where rats achieve maximal load when volitional failure is reached. This schema has not yet been described in rodent studies of maximal strength and offers a standardized and replicable method to evaluate maximal strength that can be used not only in studies of BFR-E, but also more broadly in the study of resistance exercise. In the setting of BFR-E, our data suggest that our model of BFR-E in rats leads to increased maximal strength compared to pre-training loads.

Studies that offer intramuscular insight to BFR-E physiology are limited(39,40). Further, existing studies investigating intramuscular effects of BFR-E do not isolate individual muscles by functional property. It is well known that the quadriceps include four distinct muscles, each with a unique composition of muscle fibers that inform its function (41). The Rectus Femoris is comprised predominantly of fast or type 2 muscle fibers that rely heavily on glycolytic metabolism (42). Thus, it contributes to movements that require more intense, short-lived contractile output. The VI is mostly composed of slow or type 1 muscle fibers that use oxidative metabolism to produce energy substrates (42). The Vastus Intermedius contributes to activities that involve less intense, more sustained contractile force. This can be visually confirmed: the RF appears paler, and the VI appears more red due to a low and high abundance of myoglobin, respectively.

In this study, we isolated the RF and VI to investigate the differential response to BFR-E in glycolytic and oxidative skeletal muscle. Quantifying intramuscular *HIF1a* gene expression in each muscle type, we examined the intramuscular response to BFR-E-induced oxygen deprivation. We observed muscle-specific effects of chronic BFR-E on *Hif1a* gene expression, wherein *Hif1a* was significantly increased in the VI, but not the RF. These data are consistent with existing studies of vascular responses to BFR-E. Ferguson et al. similarly observed an increase in mRNA expression of Hif1a in humans following BFR-E (43); however, muscle-specific quantifications were not performed. Lactate is well-documented to play several roles in working skeletal muscle, including as a marker of metabolic stress. We observed a similar trend, wherein intramuscular lactate was increased in the VI, but not the RF, after chronic BFR-E exposure. Accumulation of these markers not only suggests that our novel model of BFR-E in rats induces a cellular response to external vascular restriction, but it also could suggest that oxidative-predominant muscles are more responsive and/or more severely impacted by oxygen deprivation than glycolytic-predominant muscles. Whether the upregulation of mTOR gene expression observed in the RF of BFR-E rats is an indication of compensation by the RF to drive adaptation is unknown but could explain this dynamic.

When evaluating whether this differential response was sustained in genes related to the phenotypic profile of each muscle type, we observed that the VI increases gene expression of oxidative genes in BFR-E rats. Similarly, expression of glycolytic genes increased in the RF of BFR-E rats. These data suggest that BFR-E may enhance behaviors inherent to each muscle type, wherein oxidative and glycolytic muscles display increased oxidative and glycolytic behavior, respectively. Increases in these genes are consistent with other studies of glycolytic and oxidative metabolism following BFR-E (44), but have not been evaluated in a muscle-specific manner.

Our data suggest structural evidence of muscle hypertrophy following BFR-E in, to our knowledge, the first validated *in vivo* rat model of BFR-E. Although no statistical significance exists, we observed an encouraging pattern that suggests the model may lead to an increase in muscle fiber size in the BFR-E animals. These findings are consistent with the literature describing structural changes following RE. Al-Sarraf et al. demonstrate that rats subjected to RE via weight-lifting using a novel pulley system show increased muscle fiber area and an increased proportion of fast fibers compared to sedentary and treadmill-exposed rats (45). However, while we did not observe a change in the proportion of fast and slow muscle fibers within the quadriceps of BFR-E rats, our data demonstrate a muscle-specific upregulation of genes encoding fast fiber structural proteins, which could indicate increased recruitment of type 2 fibers.

Together, these molecular findings offer an alternate perspective to discussions surrounding the theoretical mechanisms of BFR-E. Proposed ideas posit that BFRE shifts muscle to a more fast-like phenotype, wherein type 2 fibers become more “type 2-like” and type 1 fibers transition to a more type 2 phenotype (12). This is thought to occur in response to several preceding molecular changes, like upregulation of systemic and intramuscular metabolites, increased mechanical tension, and activated protein signaling pathways (12,15). The data presented here provides further insight into these theories and suggests an alternative view. While we do observe an upregulation of fast fiber gene expression, this upregulation is limited to glycolytic muscles. Molecularly, the data indicate that rather than the skeletal muscle broadly transitioning to a fast-like phenotype, we see glycolytic and oxidative muscles may be becoming more enhanced versions of themselves in response to BFR-E. This shift in perspective is also demonstrated structurally. Previous theories lead us to expect to see a higher proportion of type 2 fibers in muscles of BFR-E rats; however, we report a similar proportion of type 2 fibers when comparing sedentary, exercised, and BFR-E groups.

To understand BFR-E physiology in the rehabilitative setting, we conducted a crossover design, clinical study of acute BFR-E in patients recovering from ACL-R. The study involved two visits where participants served as their own control. ACL-R and control groups were well balanced with no appreciable difference in demographics or qualitative metrics. As a preliminary analysis, metabolites were quantified from ACL and healthy control participants only after the BFR-E visit. The acute changes observed in rat plasma are not reflected in the human metabolomic analysis, however, this could be due to inherent limitations of these data. Without data from the non-BFR-E visit of each group, the statistical power is limited, which could hide group-dependent differences in the acute BFR-E response. Further, physiologic variability and small sample sizes likely contribute to the lack of significant changes observed, however it may not indicate a lack of physiologic change. Further exploration is warranted into the accurate characterization of acute BFR-E in humans.

While these *in vivo* and human data do not provide direct mechanisms underlying BFR-E-driven adaptations, it offers the foundation for several critical insights. First, our metabolomic probe in plasma of acute and chronic exercised rats reveals that BFR-E may induce a greater stimulus on the circulating metabolome in the acute condition compared to the chronic. We report significant alterations to circulating metabolites related to glucose-6-phosphate, palmitic and oleic acids, and glutathione in response to acute-BFR-E, whereas alterations are sustained in glutamic acid and malic acid following chronic BFR-E. It’s possible that these data reveal a time-dependent tolerability or familiarization with repeated BFR-E exposure, with circulating metabolic markers returning to a physiologic baseline over time. These data also suggest a potential shift in the landscape of glucose utilization in the acute setting. This would need to be evaluated intramuscularly but could be rationalized by an accelerated fatiguing and subsequent reduced functioning of type 1 muscle fibers, thereby increasing the contribution of type 2 fibers in the acute setting. While we found that the acute changes observed in rat plasma may reflect metabolomic responses to acute BFR-E in humans, we anticipate improved interpretation of this dynamic with a larger sample size.

Limitations of this study include: (1) Data are presented from a small sample size of five animals per group in the rodent study of BFR-E. Small sample sizes limit the generalizability of these findings, and trends within the data may be obscured by high intra-group variability. (2) Molecular changes observed in the study are highly dependent on genomic analysis. Central dogma dictates that genomic findings may not reflect physiology at the protein or phenotypic level. Here, we gain insight into genomic and broad functional change, but further analysis of protein-level expression changes will provide more comprehensive insight into the BFR-E effects observed.

Collectively, our data redefine the physiologic basis of BFR-E by demonstrating that robust functional and intramuscular adaptations can occur in the absence of detectable acute systemic responses in humans. This work shifts the field away from models centered on transient circulating factors and toward mechanisms rooted in muscle-specific and cumulative adaptations. As such, these findings establish a translational framework to guide the therapeutic application and mechanistic interrogation of BFR-E.

## Data Availability

All data produced in the present work are contained in the figures. Should readers be interested in raw numbers, these are available upon reasonable request to the authors.

## ACKNOWLEDGMENTS

The authors thank several contributors to this work: Dr. Wanling Zhu for her immense surgical expertise; Yale Comparative Medicine Research Histology Core for assistance with tissue embedding, slide preparation, and staining; the Cell Preparation and Analysis Core within the Yale Cooperative Center of Excellence in Hematology for gracious access to the Hemavet analyzer; Dr. Elena Gracheva for use of the EchoMRI Body Composition Analyzer; and the clinical teams at Yale New Haven Health Orthopedic and Rehabilitation Clinic and Gaylord Physical therapy clinic for facilitation and support of the clinical study. This publication was made possible by CTSA Grant Number UL1 TR001863 from the National Center for Advancing Translational Science (NCATS), a component of the National Institutes of Health (NIH), and a supplement to NIH R37CA258261-A1. Its contents are solely the responsibility of the authors and do not necessarily represent the official views of NIH.

## AUTHOR CONTRIBUTIONS

Conceptualization, A.E.F and R.J.P.; Methodology, A.E.F, S.C.B.R.N., C.A., C.T.; Investigation, A.E.F., S.C.B.R.N., M.D., Z.L. C.P.; Writing, Review & Editing, A.E.F, C.T. and R.J.P.; Funding Acquisition, R.J.P.; Resources, R.J.P.; Supervision, R.J.P. and C.A.

## Supplemental Tables And Figures

**Supplemental Table 1.**
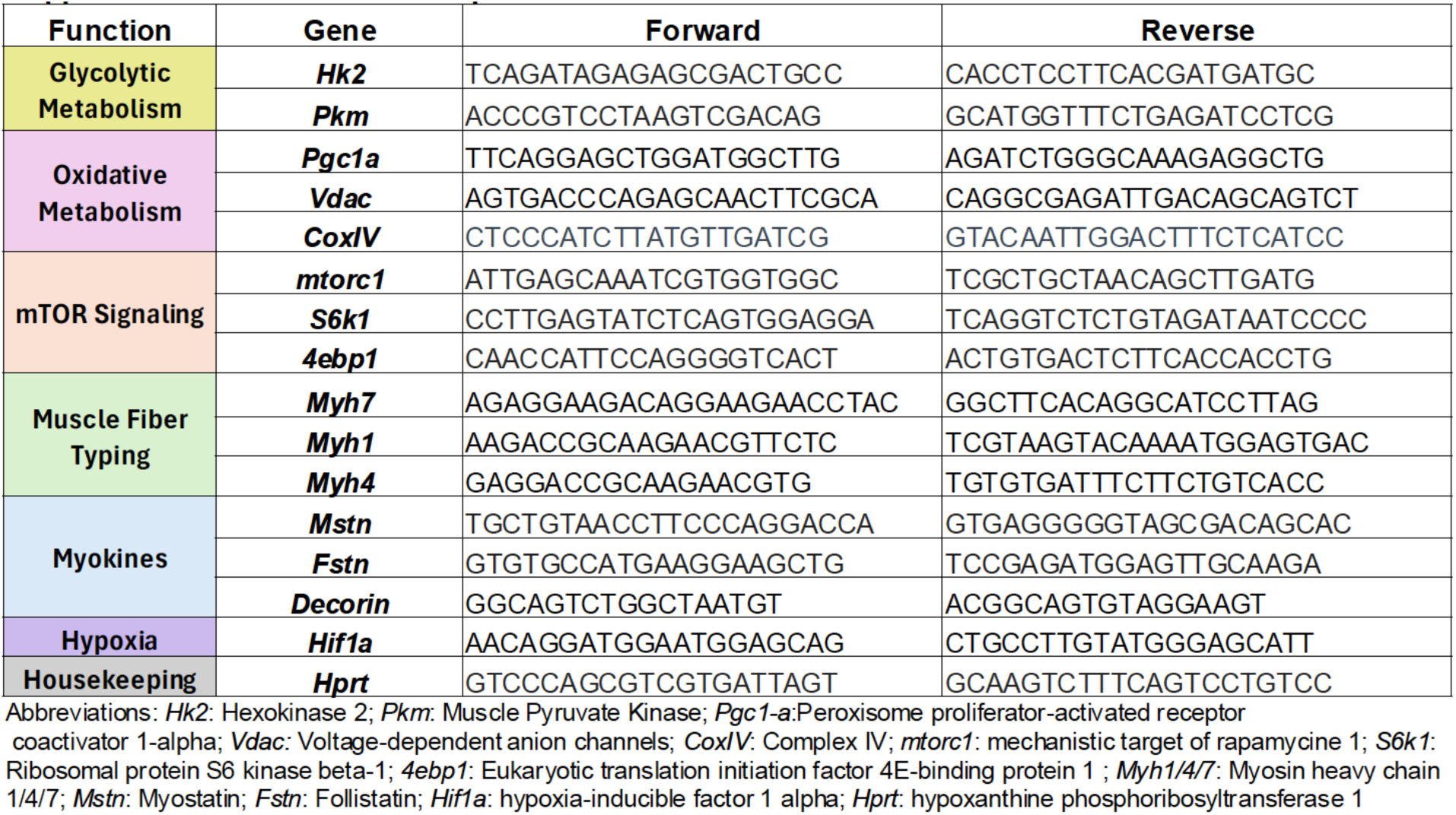
Quantitative Polymerase Chain Reaction Primer Sequences. Forward and Reverse primer sequences for quantification of genes related to glycolytic metabolism, oxidative metabolism, mTOR signaling, muscle fiber typing, myokines, and hypoxia.

**Supplemental Table 2.**
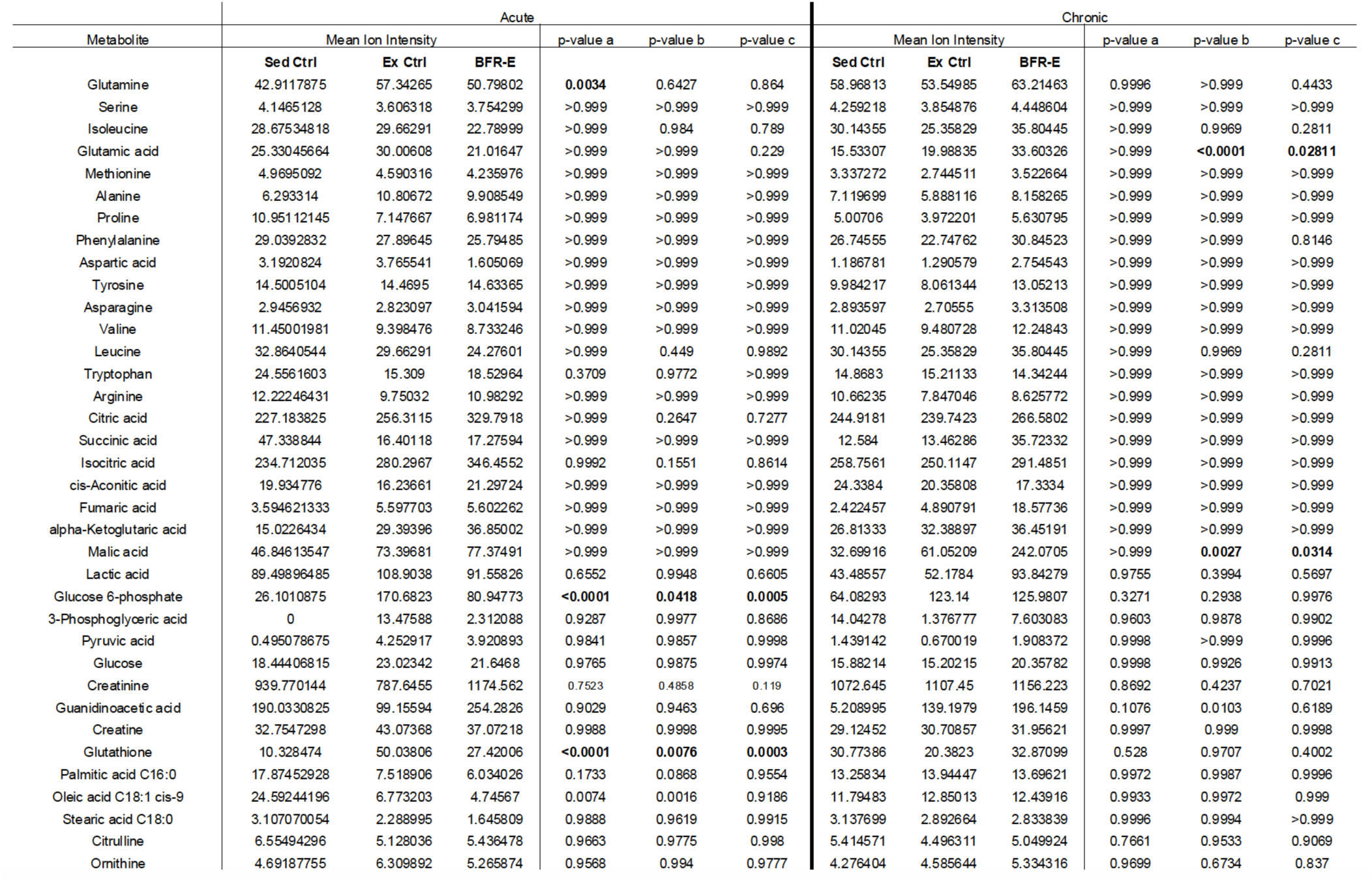
Quantitative Mean Ion Intensities Following Metabolomic Analyses in Rats. Metabolite ion intensities identified and quantified using LC-MS from plasma isolated from sedentary control (Sed Ctrl), exercise control (Ex ctrl), and blood flow restriction exercise (BFR-E) rats exposed to either acute or chronic resistance exercise. P-value a icompares sedentary control vs exercise control; p-value b compares sedentary control vs BFR-E; p value c compares exercise control vs BFR-E. Significance set to p<0.05.

**Supplemental Figure 1.**
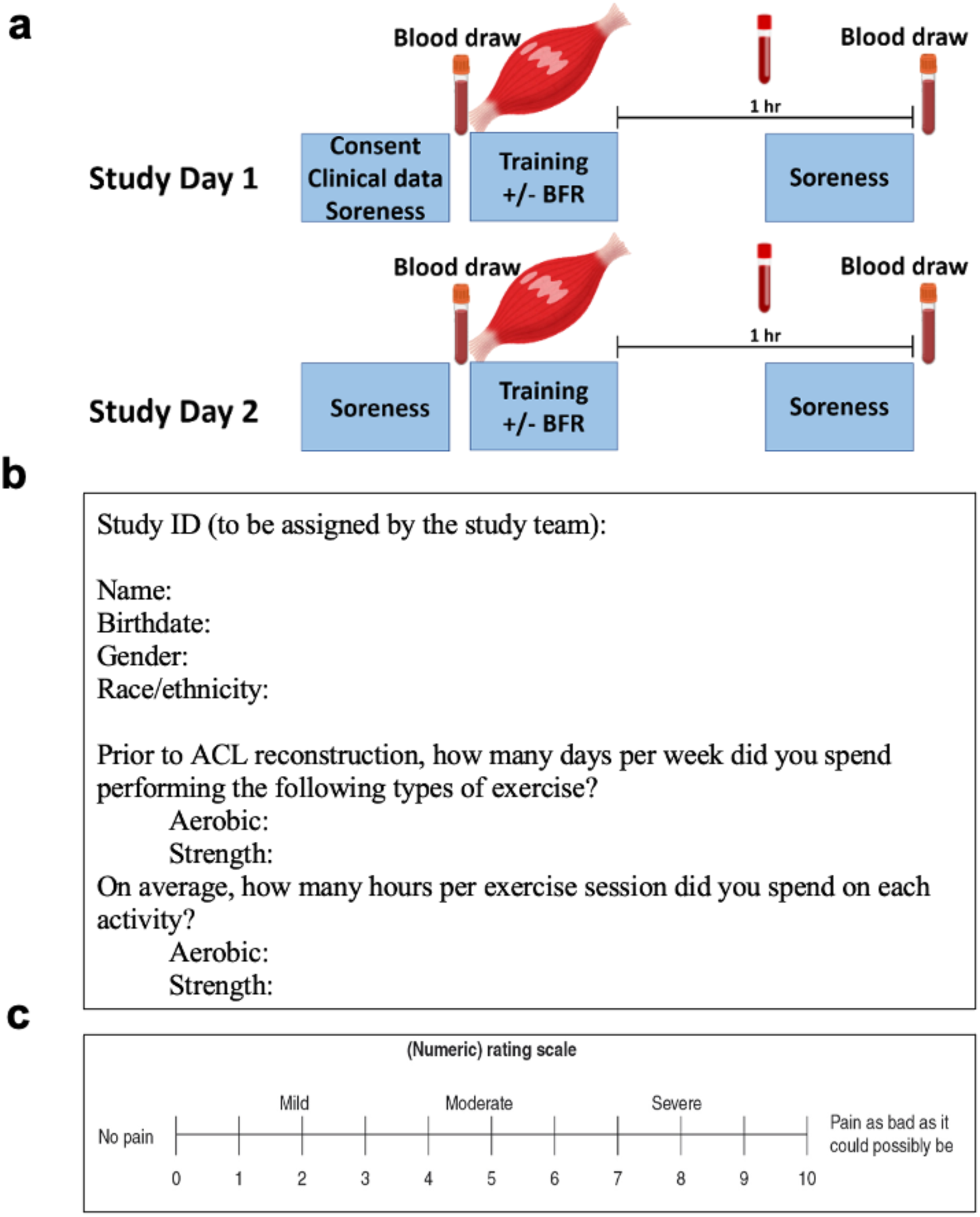
Clinical Study Schematic. (a) Each participant underwent two study visits during which they completed qualitative surveys, single-leg knee extension exercise with or without BFR, and pre- and post-exercise blood sampling. (b) Activity survey used to assess baseline exercise behavior. (c) Representative diagram of soreness scale used to determine perceived pain before and after exercise session during each visit. Abbreviations: BFR = blood flow restriction; ACL = anterior cruciate ligament

**Supplemental Figure 2.**
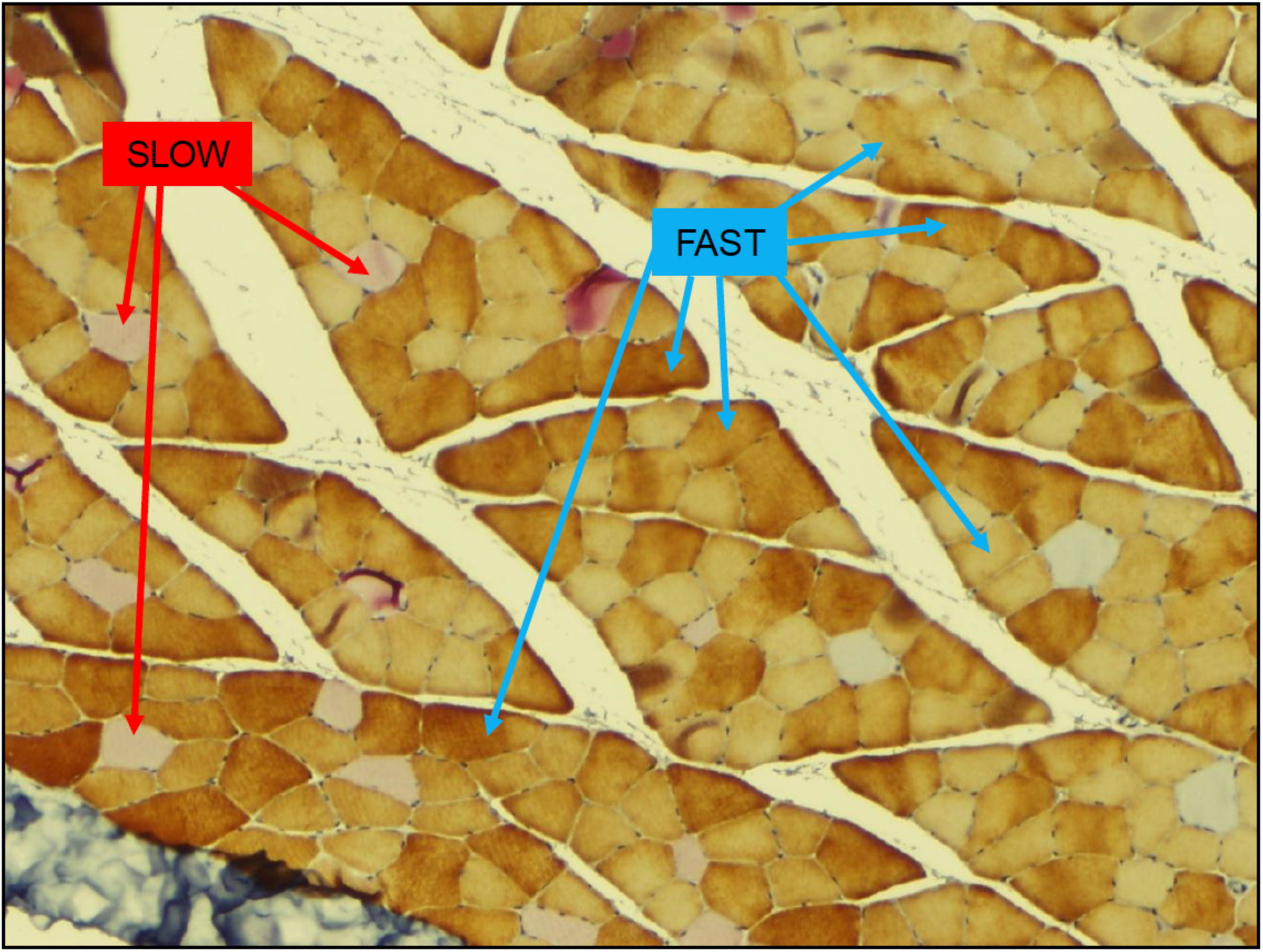
Muscle Fiber Type Identification. Immunohistochemistry of quadriceps muscle cut in cross-section and double stained with fast and slow myosin. Slow and fast fibers identified as fibers stained pink (red arrows) and brown (blue arrows), respectively.

